# Geographic and clinical predictors of one-day discharge after radical prostatectomy: a SEER-Medicare cohort study

**DOI:** 10.64898/2026.09.16.26363208

**Authors:** Jacob Hanna, Megan R. Shanahan, Jennifer Sykes, Farzin Khosrow-Khavar, Thomas L. Jang, Hari S. Iyer

## Abstract

**Objectives:** To study the geographic patterns, temporal trends, and predictors of one-day discharge following radical prostatectomy.

**Methods:** We conducted a retrospective cohort study of men diagnosed with prostate cancer from 2008 to 2019 in the Surveillance, Epidemiology, and End Results (SEER)-Medicare linked database. Patients with metastatic disease, health maintenance organization coverage, or age younger than 66 years were excluded. Type of surgery and length of stay were derived from inpatient claims. We compared one-day discharge by early (2008–2013) and late (2014–2019) periods, by census region, and by individual- and facility-level factors, and used generalized estimating equations to estimate adjusted odds ratios (aOR) with 95% confidence intervals (CI), accounting for clustering of patients within hospitals.

**Results:** Among 40,728 patients treated at 1,048 hospitals, 52.4% were discharged on postoperative day 1, increasing from 48.8% (2008–2013) to 56.8% (2014–2019). Higher odds of one-day discharge were associated with robotic surgery (aOR 2.88, 95% CI 2.54–3.27) and treatment at a National Cancer Institute-designated comprehensive cancer center (aOR 1.88, 95% CI 1.28–2.76). Black versus White patients (aOR 0.76, 95% CI 0.70–0.82), unmarried versus married patients (aOR 0.79, 95% CI 0.73–0.85), and patients in the lowest versus highest neighborhood socioeconomic quintile (aOR 0.86, 95% CI 0.79–0.94) had lower odds. Odds were 31% higher in the late versus early period (aOR 1.31, 95% CI 1.20–1.43). Regional differences were attenuated in sensitivity analyses.

**Conclusions:** One-day discharge after radical prostatectomy has increased over time and is now the predominant practice among Medicare beneficiaries, driven primarily by robotic surgery and treatment era. Persistent racial and socioeconomic differences in discharge patterns warrant further research into whether early discharge is equally safe and beneficial across patient populations.

## Introduction

The incidence of prostate cancer is rising, with 333,830 cases estimated for 2026 [1]. Radical prostatectomy is an important first-line treatment option for men with clinically localized prostate cancer [2]. While no formal Enhanced Recovery After Surgery (ERAS) protocols exist specifically for radical prostatectomy, institutional adaptations of general ERAS principles have been studied and implemented since at least the early 2010s [3].

Robot-assisted radical prostatectomy offers several advantages over the open approach, including lower blood loss and shorter hospital stay [4–7]. These surgical advances, combined with institutional adoption of ERAS principles, have improved perioperative recovery [8–11]. By 2009, minimally invasive prostatectomy had overtaken open surgical approaches as the dominant technique worldwide [12,13].

One-day discharge (ODD), defined as a length of stay of one calendar day with discharge on postoperative day 1, has emerged as a marker of efficient perioperative care. Few studies have examined the predictors of ODD after radical prostatectomy or assessed regional variation in its adoption. Previous research documented rapid centralization of prostatectomy during early robotic adoption, with hospitals offering robotic surgery performing 85% of procedures by 2009, despite representing only 35% of treating facilities [12]. Whether this concentration of surgical volume has translated into geographic disparities in discharge outcomes remains unclear.

Using Surveillance, Epidemiology, and End Results (SEER)-Medicare data from 2008 to 2019, we sought to identify predictors of ODD after radical prostatectomy and to examine geographic and temporal trends in the adoption of ODD.

## Materials and methods

### Study population and design

We conducted a retrospective cohort study using the SEER-Medicare linked database, which combines population-based cancer registry data with Medicare claims for beneficiaries aged 65 years and older. The SEER-Medicare database includes detailed clinical cancer information from 20 SEER registries and provides a unique resource for the longitudinal tracking of care delivery patterns across diverse healthcare settings [14]. This study is reported in accordance with the Strengthening the Reporting of Observational Studies in Epidemiology (STROBE) guidelines (S1 Checklist). Data were accessed for research purposes between August 1, 2025 and June 4 2026. Authors did not have access to information that could identify individual participants during or after data collection.

Men diagnosed with prostate cancer between 2008 and 2019 were identified from the SEER-Medicare linked database, yielding 784,790 patients. Of these, 98,388 underwent radical prostatectomy, as identified through the ICD procedure codes specified in S1 Table. After excluding patients younger than 66 years (n = 24,776) to ensure an adequate lookback period for comorbidity assessment, 73,612 Medicare-eligible patients remained. We then excluded patients with Health Maintenance Organization (HMO) enrollment or without continuous fee-for-service Medicare Part A and B coverage (n = 29,790), leaving 43,822 eligible patients. Additional exclusion criteria for metastatic disease (n = 2,977), in situ or distant stage disease (n = 114), and missing hospital data (n = 3) yielded a final analytic cohort of 40,728 patients treated across 1,048 hospitals (Fig 1).

**Fig 1.**
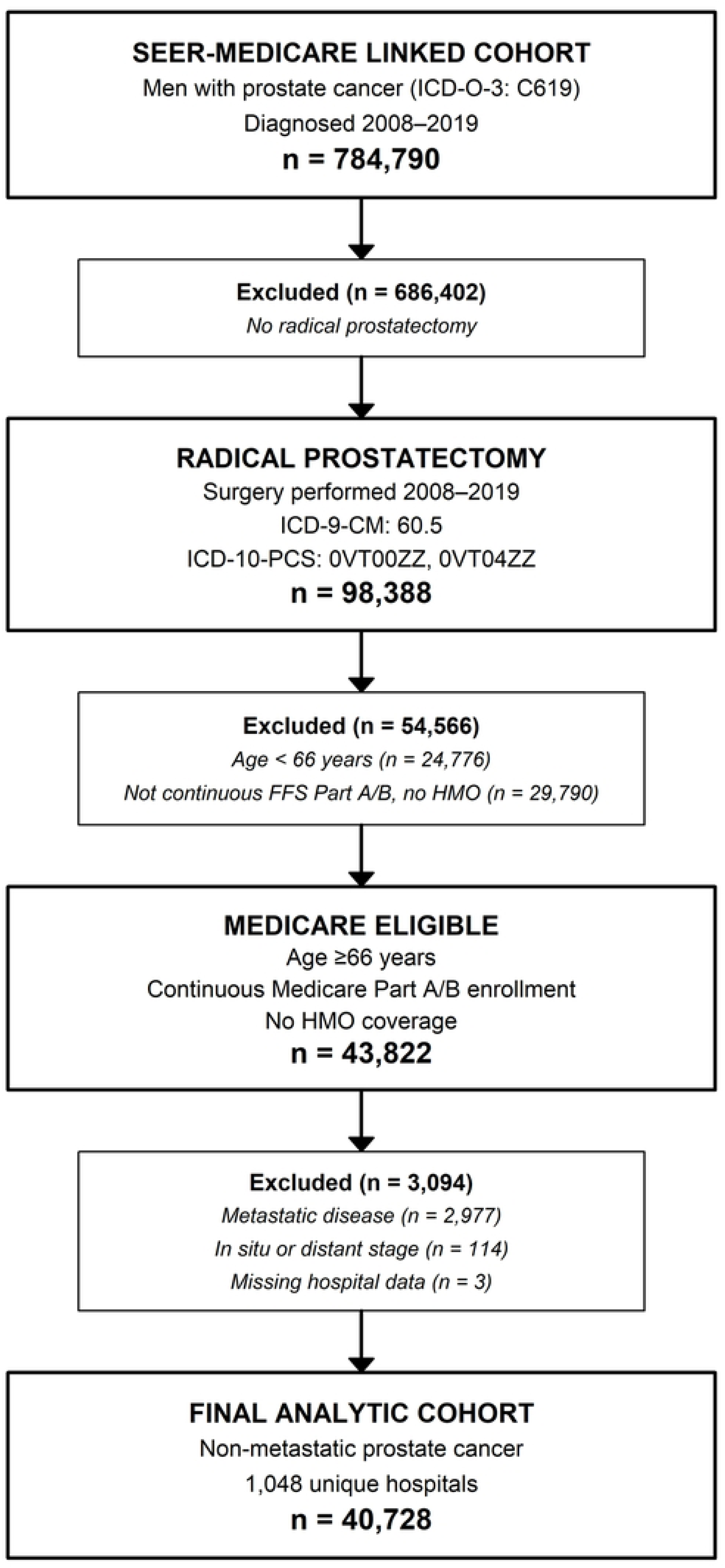
Study cohort selection. SEER-Medicare patients diagnosed with prostate cancer (2008– 2019) who underwent radical prostatectomy were identified. Sequential exclusion criteria were applied to arrive at the final analytic cohort of 40,728 patients treated across 1,048 hospitals. SEER, Surveillance, Epidemiology, and End Results; HMO, Health Maintenance Organization.

### Ethics statement

This study was approved by the Rutgers University Institutional Review Board (Study ID Pro2021001673) and reviewed by the National Cancer Institute SEER-Medicare program for adherence to the data use agreement. The requirement for informed consent was waived because the study used de-identified secondary data. Data were accessed for research purposes under the SEER-Medicare data use agreement; the authors did not have access to information that could identify individual participants.

### Measures

Robotic assistance was identified from the MEDPAR inpatient procedure fields using procedure codes detailed in S1 Table, with robotic procedures representing 67.4% of cases. ODD was defined as a length of stay of one calendar day from hospital admission to discharge, corresponding to discharge on postoperative day 1, based on the LOS_DAY_CNT variable in the MEDPAR inpatient files. All ODD events in our cohort represent an overnight inpatient stay. Because the cohort was identified from inpatient claims, same-day discharges performed as outpatient procedures, which have become increasingly feasible with single-port robotic platforms, were not included in this analysis. For patients with multiple claims for radical prostatectomy, only the first record was retained.

### Covariates

Demographic and clinical variables were obtained from the SEER cancer registry files. These included age at diagnosis (66–69, 70–74, 75–79, 80+ years), race/ethnicity (Non-Hispanic White, Non-Hispanic Black, Hispanic, and Other/Unknown), marital status (married, not married, and unknown), SEER summary cancer stage (localized, regional, or unknown) [15], Gleason score (≤6, 7, 8–10, or missing), and prostate-specific antigen (PSA) level at diagnosis (≤10, 10–20, >20 ng/mL, or missing). Census tract neighborhood socioeconomic status (SES) at diagnosis was calculated using the Yost index [16,17] and modeled as quintiles from the lowest (disadvantaged) to the highest (advantaged). Rural/urban residence was classified as metropolitan or non-metropolitan. The Klabunde-Charlson Comorbidity Index (CCI) was calculated using inpatient diagnosis codes from MEDPAR and outpatient diagnosis codes from Medicare Outpatient files during the 12 months prior to admission (S2 Table) to ensure comprehensive comorbidity capture. CCI scores were categorized as 0, 1, and 2+ [18]. Hospital characteristics were obtained from the SEER-Medicare Hospital File, including size (large ≥400 beds, medium 200–399 beds, small <200 beds), teaching status (yes/no), National Cancer Institute (NCI) designation (non-NCI, NCI clinical, NCI comprehensive), and Commission on Cancer (CoC) accreditation (yes/no).

The SEER registries were grouped into four U.S. census regions based on patient residence. The 20 registries were distributed as follows: West region (California, Idaho, New Mexico, Utah, Washington; 8 registries), Northeast region (Connecticut, Massachusetts, New Jersey, New York; 4 registries), South region (Georgia, Kentucky, Louisiana, Texas; 6 registries), and Midwest region (Iowa, Michigan; 2 registries), with the Northeast serving as the reference group. Temporal trends were evaluated by dividing the study period into early (2008–2013) and late (2014–2019) periods to assess innovation diffusion patterns.

### Statistical analysis

We used generalized estimating equations (GEE) with a logistic link to model ODD as a binary outcome, using an exchangeable correlation structure (correlation = 0.21) and empirical standard errors to account for the clustering of patients within 1,048 hospitals. A univariable unadjusted GEE model was first fit for each covariate individually, followed by a multivariable-adjusted model including patient-level covariates (age category, race/ethnicity, marital status, SES quintile, rural/urban residence, cancer stage, CCI, Gleason score, PSA category, and surgical approach), temporal controls (era), geographic controls (SEER region), and hospital-level covariates (hospital size, teaching status, NCI designation, and CoC accreditation).

All variables were selected a priori based on their clinical relevance. Variables with missing data (Gleason score, PSA, cancer stage, teaching hospital status, SES quintile, and NCI/CoC designation) were modeled with missing indicators to retain the full analytic sample. A sensitivity analysis was conducted by re-estimating the multivariable GEE model after excluding Gleason score, PSA, and cancer stage, which were variables with substantial missing data, to assess whether missingness influenced regional estimates. All statistical analyses were conducted using SAS version 9.4 (SAS Institute, Cary, NC), with significance defined as a two-sided P < 0.05.

## Results

The final analytic cohort included 40,728 patients who underwent radical prostatectomy between 2008 and 2019 across 1,048 hospitals (Table 1). The mean age was 69.8 (SD 3.1) years, with 83.1% Non-Hispanic White, 6.4% Non-Hispanic Black, 7.1% Hispanic, and 2.9% Asian American/Native Hawaiian/Pacific Islander. Overall, 21,344 patients (52.4%) had an ODD. Hospital characteristics varied substantially across regions (Table 2). Most patients were treated at large hospitals (≥400 beds; 57%) and teaching hospitals (69%). Twenty percent of patients were treated at NCI-designated cancer centers, with 15% treated at comprehensive centers and 5% at clinical centers.

**Table 1.** Patient and clinical characteristics by geographic region, radical prostatectomy patients, SEER-Medicare 2008–2019 (N = 40,728).

| Characteristic | Overall<br>N = 40,728 | Geographic region |  |  |  |
| --- | --- | --- | --- | --- | --- |
|  |  | Northeast<br>N = 10,841 | South<br>N = 13,240 | West<br>N = 13,710 | Midwest<br>N = 2,937 |
| Age Category |  |  |  |  |  |
| 66-69 | 21,588 (53.0) | 6,219 (57.4) | 7,218 (54.5) | 6,485 (47.3) | 1,666 (56.7) |
| 70-74 | 15,091 (37.1) | 3,755 (34.6) | 4,822 (36.4) | 5,442 (39.7) | 1,072 (36.5) |
| 75-79 | 3,445 (8.5) | 712 (6.6) | 1,036 (7.8) | 1,520 (11.1) | 177 (6.0) |
| 80+ | 604 (1.5) | 155 (1.4) | 164 (1.2) | 263 (1.9) | 22 (0.7) |
| Race/Ethnicity |  |  |  |  |  |
| AANHPI | 1,163 (2.9) | 238 (2.2) | 179 (1.4) | 724 (5.3) | 22 (0.7) |
| Hispanic | 2,905 (7.1) | 591 (5.5) | 880 (6.6) | 1,409 (10.3) | 25 (0.9) |
| NH-Black | 2,596 (6.4) | 756 (7.0) | 1,177 (8.9) | 371 (2.7) | 292 (9.9) |
| NH-White | 33,860 (83.1) | 9,176 (84.6) | 10,987 (83.0) | 11,109 (81.0) | 2,588 (88.1) |
| Missing | 204 (0.5) | 80 (0.7) | 17 (0.1) | 97 (0.7) | 10 (0.3) |
| Marital Status |  |  |  |  |  |
| Married | 19,493 (47.9) | 2,797 (25.8) | 4,278 (32.3) | 10,133 (73.9) | 2,285 (77.8) |
| Not Married | 3,960 (9.7) | 581 (5.4) | 819 (6.2) | 2,108 (15.4) | 452 (15.4) |
| Unknown | 17,275 (42.4) | 7,463 (68.8) | 8,143 (61.5) | 1,469 (10.7) | 200 (6.8) |
| Gleason Score |  |  |  |  |  |
| ≤6 | 5,792 (14.2) | 926 (8.5) | 1,388 (10.5) | 2,905 (21.2) | 573 (19.5) |
| 7 | 10,024 (24.6) | 1,475 (13.6) | 2,208 (16.7) | 5,017 (36.6) | 1,324 (45.1) |
| 8-10 | 4,335 (10.6) | 721 (6.7) | 830 (6.3) | 2,281 (16.6) | 503 (17.1) |
| Missing | 20,577 (50.5) | 7,719 (71.2) | 8,814 (66.6) | 3,507 (25.6) | 537 (18.3) |
| PSA Category |  |  |  |  |  |
| 10–20 | 3,288 (8.1) | 345 (3.2) | 652 (4.9) | 1,835 (13.4) | 456 (15.5) |
| ≤10 | 17,632 (43.3) | 2,735 (25.2) | 3,866 (29.2) | 8,992 (65.6) | 2,039 (69.4) |
| >20 | 1,133 (2.8) | 145 (1.3) | 239 (1.8) | 606 (4.4) | 143 (4.9) |
| Missing | 18,675 (45.9) | 7,616 (70.3) | 8,483 (64.1) | 2,277 (16.6) | 299 (10.2) |
| Disease Stage |  |  |  |  |  |
| Localized | 26,409 (64.8) | 7,044 (65.0) | 8,847 (66.8) | 8,748 (63.8) | 1,770 (60.3) |
| Regional | 13,786 (33.8) | 3,672 (33.9) | 4,075 (30.8) | 4,876 (35.6) | 1,163 (39.6) |
| Missing | 533 (1.3) | 125 (1.2) | 318 (2.4) | 86 (0.6) | 4 (0.1) |
| Charlson Comorbidity |  |  |  |  |  |
| 0 | 24,870 (61.1) | 6,503 (60.0) | 7,970 (60.2) | 8,592 (62.7) | 1,805 (61.5) |
| 1 | 9,641 (23.7) | 2,569 (23.7) | 3,259 (24.6) | 3,111 (22.7) | 702 (23.9) |
| 2+ | 6,217 (15.3) | 1,769 (16.3) | 2,011 (15.2) | 2,007 (14.6) | 430 (14.6) |
| Robotic Surgery |  |  |  |  |  |
| Non-robotic | 13,289 (32.6) | 3,524 (32.5) | 4,485 (33.9) | 4,366 (31.8) | 914 (31.1) |
| Robotic | 27,439 (67.4) | 7,317 (67.5) | 8,755 (66.1) | 9,344 (68.2) | 2,023 (68.9) |
|  |  | Northeast<br>N = 10,841 | South<br>N = 13,240 | West<br>N = 13,710 | Midwest<br>N = 2,937 |
| Treatment Era |  |  |  |  |  |
| Early | 23,634 (58.0) | 5,938 (54.8) | 7,495 (56.6) | 8,443 (61.6) | 1,758 (59.9) |
| Late | 17,094 (42.0) | 4,903 (45.2) | 5,745 (43.4) | 5,267 (38.4) | 1,179 (40.1) |
| SES Quintile |  |  |  |  |  |
| Q1 | 4,266 (10.5) | 493 (4.5) | 2,486 (18.8) | 969 (7.1) | 318 (10.8) |
| Q2 | 5,776 (14.2) | 838 (7.7) | 2,739 (20.7) | 1,690 (12.3) | 509 (17.3) |
| Q3 | 7,182 (17.6) | 1,359 (12.5) | 2,662 (20.1) | 2,374 (17.3) | 787 (26.8) |
| Q4 | 8,962 (22.0) | 2,739 (25.3) | 2,342 (17.7) | 3,150 (23.0) | 731 (24.9) |
| Q5 | 13,414 (32.9) | 4,843 (44.7) | 2,748 (20.8) | 5,335 (38.9) | 488 (16.6) |
| Missing | 1,128 (2.8) | 569 (5.2) | 263 (2.0) | 192 (1.4) | 104 (3.5) |
| One-Day Discharge |  |  |  |  |  |
| No | 19,384 (47.6) | 4,691 (43.3) | 6,517 (49.2) | 6,680 (48.7) | 1,496 (50.9) |
| Yes | 21,344 (52.4) | 6,150 (56.7) | 6,723 (50.8) | 7,030 (51.3) | 1,441 (49.1) |

Data are n (%) unless noted. AANHPI, Asian American/Native Hawaiian/Pacific Islander; NH, Non-Hispanic; PSA, prostate-specific antigen; Q, quintile; SES, socioeconomic status (Yost index).

**Table 2.** Hospital characteristics and one-day discharge by geographic region, radical prostatectomy patients, SEER-Medicare 2008–2019 (N = 40,728).

| Characteristic | Overall<br>N = 40,728 | Geographic region |  |  |  |
| --- | --- | --- | --- | --- | --- |
|  |  | Northeast<br>N = 10,841 | South<br>N = 13,240 | West<br>N = 13,710 | Midwest<br>N = 2,937 |
| Hospital Bed Size |  |  |  |  |  |
| Large (≥400) | 23,135 (56.8) | 7,125 (65.7) | 7,706 (58.2) | 6,363 (46.4) | 1,941 (66.1) |
| Medium (200–399) | 11,830 (29.0) | 2,723 (25.1) | 2,725 (20.6) | 5,649 (41.2) | 733 (25.0) |
| Small (<200) | 5,763 (14.1) | 993 (9.2) | 2,809 (21.2) | 1,698 (12.4) | 263 (9.0) |
| Hospital Ownership |  |  |  |  |  |
| For-Profit | 4,630 (11.4) | 370 (3.4) | 2,695 (20.4) | 1,145 (8.4) | 420 (14.3) |
| Government | 6,296 (15.5) | 537 (5.0) | 3,164 (23.9) | 2,337 (17.0) | 258 (8.8) |
| Non-Profit | 29,802 (73.2) | 9,934 (91.6) | 7,381 (55.7) | 10,228 (74.6) | 2,259 (76.9) |
| Teaching Hospital |  |  |  |  |  |
| Non-Teaching | 11,370 (27.9) | 1,264 (11.7) | 5,437 (41.1) | 4,459 (32.5) | 210 (7.2) |
| Teaching | 27,939 (68.6) | 9,436 (87.0) | 7,248 (54.7) | 8,558 (62.4) | 2,697 (91.8) |
| Missing | 1,419 (3.5) | 141 (1.3) | 555 (4.2) | 693 (5.1) | 30 (1.0) |
| NCI Cancer Center |  |  |  |  |  |
| Non-NCI | 32,563 (80.0) | 7,784 (71.8) | 11,696 (88.3) | 10,698 (78.0) | 2,385 (81.2) |
| Clinical Cancer Center | 2,121 (5.2) | 1,196 (11.0) | 181 (1.4) | 734 (5.4) | 10 (0.3) |
| Comprehensive Cancer Center | 6,044 (14.8) | 1,861 (17.2) | 1,363 (10.3) | 2,278 (16.6) | 542 (18.5) |
| CoC Accreditation |  |  |  |  |  |
| Non-CoC | 16,730 (41.1) | 3,808 (35.1) | 5,655 (42.7) | 5,874 (42.8) | 1,393 (47.4) |
|  |  | Northeast<br>N = 10,841 | South<br>N = 13,240 | West<br>N = 13,710 | Midwest<br>N = 2,937 |
| CoC-Accredited | 23,998 (58.9) | 7,033 (64.9) | 7,585 (57.3) | 7,836 (57.2) | 1,544 (52.6) |
| **Rural/Urban** |  |  |  |  |  |
| Metro | 34,691 (85.2) | 10,229 (94.4) | 10,038 (75.8) | 12,296 (89.7) | 2,128 (72.5) |
| Non-Metro | 6,031 (14.8) | 610 (5.6) | 3,202 (24.2) | 1,410 (10.3) | 809 (27.5) |
| Unknown | 6 (0.0) | 2 (0.0) |  | 4 (0.0) |  |

Data are n (%) unless noted. CoC, Commission on Cancer; NCI, National Cancer Institute.

### Factors associated with one-day discharge

The unadjusted and adjusted odds ratios from the GEE models are presented in Table 3. The strongest predictor of ODD was robotic versus non-robotic surgery (aOR 2.88, 95% CI 2.54– 3.27). Patients treated at NCI comprehensive cancer centers (aOR 1.88, 95% CI 1.28–2.76) and NCI clinical cancer centers (aOR 1.95, 95% CI 1.09–3.50) had higher odds of ODD than those treated at non-NCI hospitals. Compared with patients aged 66–69 years, those aged 70–74 years had 8% lower odds (aOR 0.92, 95% CI 0.88–0.96), those aged 75–79 years had 29% lower odds (aOR 0.71, 95% CI 0.64–0.78), and those aged 80+ years had 72% lower odds (aOR 0.28, 95% CI 0.22–0.35). Compared with Non-Hispanic White patients, Non-Hispanic Black patients (aOR 0.76, 95% CI 0.70–0.82), Hispanic patients (aOR 0.87, 95% CI 0.80–0.95), and those of other or unknown race/ethnicity (aOR 0.84, 95% CI 0.74–0.95) had lower odds of ODD (Fig 2).

**Fig 2.**
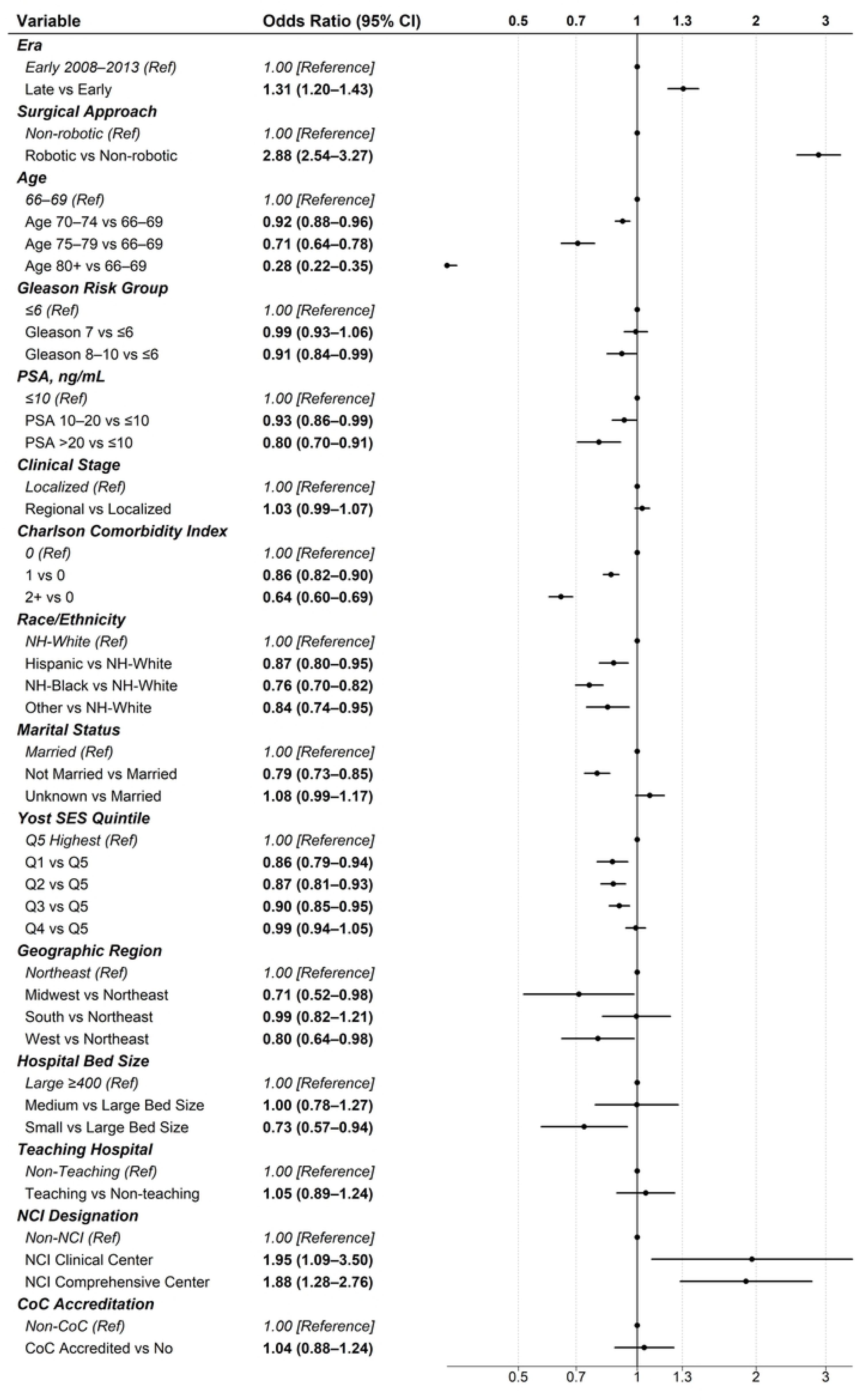
Adjusted odds ratios for one-day discharge after radical prostatectomy from the multivariable GEE model (N = 40,728). Points are adjusted odds ratios and horizontal lines are 95% confidence intervals on a log scale; the vertical line indicates an odds ratio of 1. aOR, adjusted odds ratio; CI, confidence interval; CoC, Commission on Cancer; GEE, generalized estimating equations; NCI, National Cancer Institute; NH, Non-Hispanic; PSA, prostate-specific antigen; Ref, reference; SES, socioeconomic status.

**Table 3.**
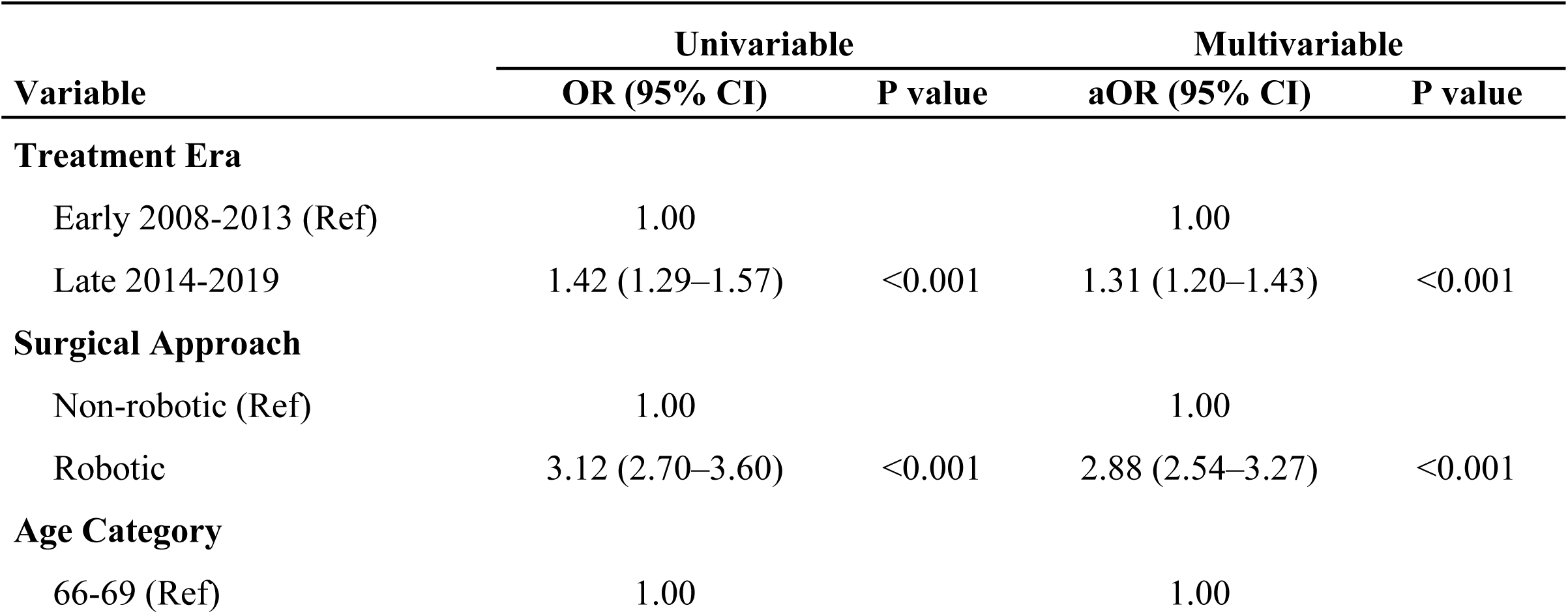
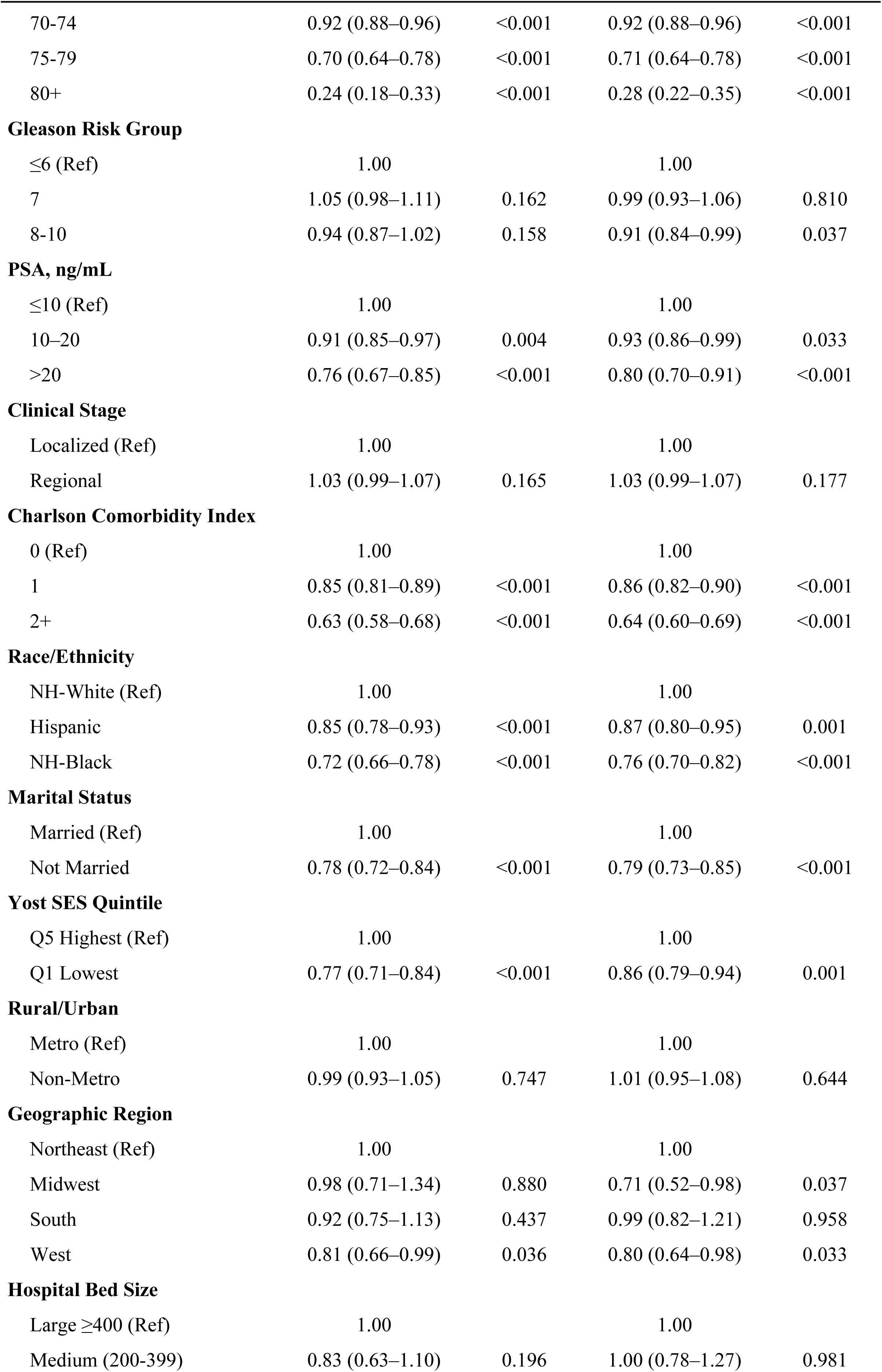
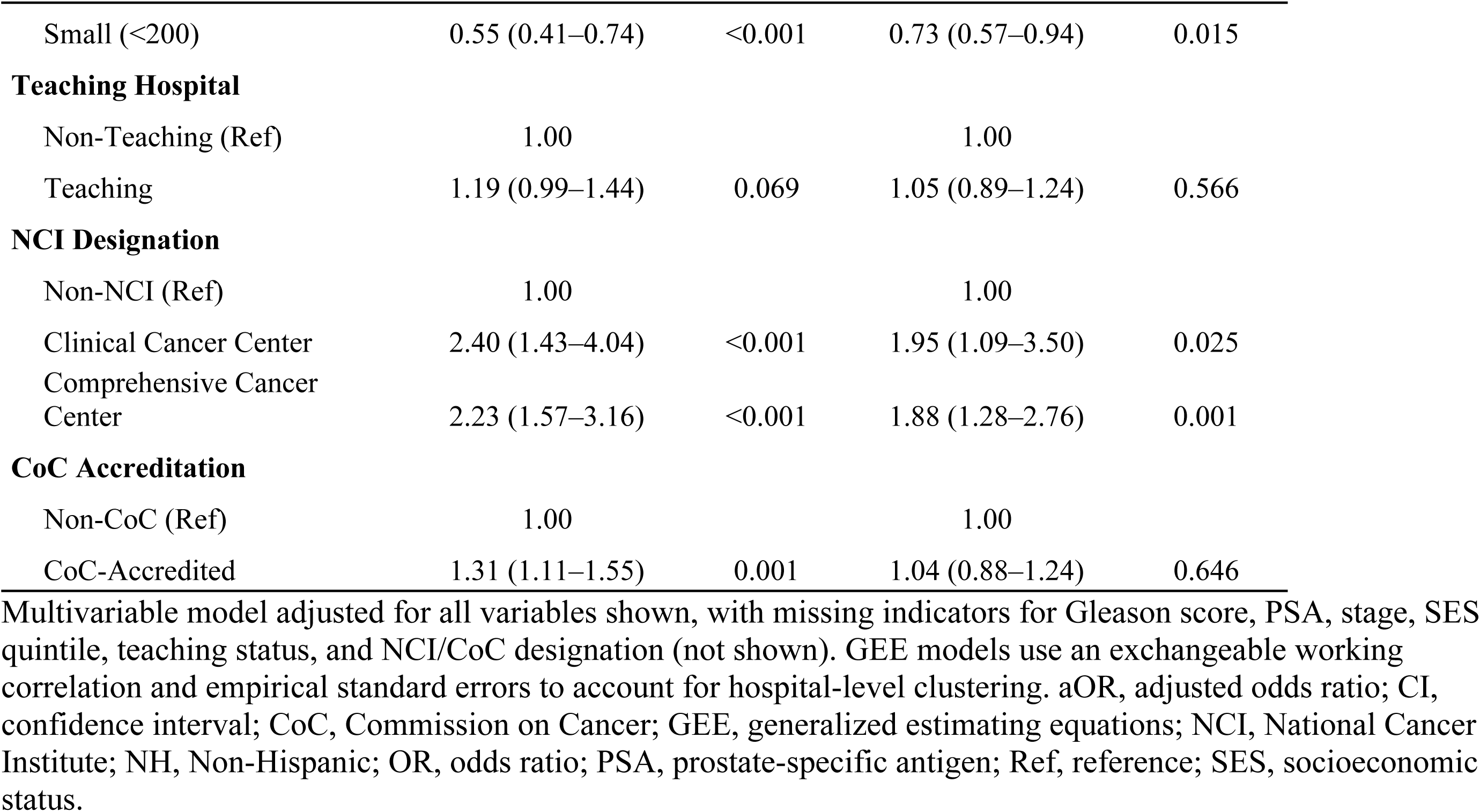
Univariable and multivariable GEE analysis of factors associated with one-day discharge after radical prostatectomy, SEER-Medicare 2008–2019 (N = 40,728).

| Variable | Univariable |  | Multivariable |  |
| --- | --- | --- | --- | --- |
|  | OR (95% CI) | P value | aOR (95% CI) | P value |
| **Treatment Era** |  |  |  |  |
| Early 2008-2013 (Ref) | 1.00 |  | 1.00 |  |
| Late 2014-2019 | 1.42 (1.29–1.57) | <0.001 | 1.31 (1.20–1.43) | <0.001 |
| **Surgical Approach** |  |  |  |  |
| Non-robotic (Ref) | 1.00 |  | 1.00 |  |
| Robotic | 3.12 (2.70–3.60) | <0.001 | 2.88 (2.54–3.27) | <0.001 |
| **Age Category** |  |  |  |  |
| 66-69 (Ref) | 1.00 |  | 1.00 |  |
|  | OR (95% CI) | P value | aOR (95% CI) | P value |
| 70-74 | 0.92 (0.88–0.96) | <0.001 | 0.92 (0.88–0.96) | <0.001 |
| 75-79 | 0.70 (0.64–0.78) | <0.001 | 0.71 (0.64–0.78) | <0.001 |
| 80+ | 0.24 (0.18–0.33) | <0.001 | 0.28 (0.22–0.35) | <0.001 |
| **Gleason Risk Group** |  |  |  |  |
| ≤6 (Ref) | 1.00 |  | 1.00 |  |
| 7 | 1.05 (0.98–1.11) | 0.162 | 0.99 (0.93–1.06) | 0.810 |
| 8-10 | 0.94 (0.87–1.02) | 0.158 | 0.91 (0.84–0.99) | 0.037 |
| **PSA, ng/mL** |  |  |  |  |
| ≤10 (Ref) | 1.00 |  | 1.00 |  |
| 10–20 | 0.91 (0.85–0.97) | 0.004 | 0.93 (0.86–0.99) | 0.033 |
| >20 | 0.76 (0.67–0.85) | <0.001 | 0.80 (0.70–0.91) | <0.001 |
| **Clinical Stage** |  |  |  |  |
| Localized (Ref) | 1.00 |  | 1.00 |  |
| Regional | 1.03 (0.99–1.07) | 0.165 | 1.03 (0.99–1.07) | 0.177 |
| **Charlson Comorbidity Index** |  |  |  |  |
| 0 (Ref) | 1.00 |  | 1.00 |  |
| 1 | 0.85 (0.81–0.89) | <0.001 | 0.86 (0.82–0.90) | <0.001 |
| 2+ | 0.63 (0.58–0.68) | <0.001 | 0.64 (0.60–0.69) | <0.001 |
| **Race/Ethnicity** |  |  |  |  |
| NH-White (Ref) | 1.00 |  | 1.00 |  |
| Hispanic | 0.85 (0.78–0.93) | <0.001 | 0.87 (0.80–0.95) | 0.001 |
| NH-Black | 0.72 (0.66–0.78) | <0.001 | 0.76 (0.70–0.82) | <0.001 |
| **Marital Status** |  |  |  |  |
| Married (Ref) | 1.00 |  | 1.00 |  |
| Not Married | 0.78 (0.72–0.84) | <0.001 | 0.79 (0.73–0.85) | <0.001 |
| **Yost SES Quintile** |  |  |  |  |
| Q5 Highest (Ref) | 1.00 |  | 1.00 |  |
| Q1 Lowest | 0.77 (0.71–0.84) | <0.001 | 0.86 (0.79–0.94) | 0.001 |
| **Rural/Urban** |  |  |  |  |
| Metro (Ref) | 1.00 |  | 1.00 |  |
| Non-Metro | 0.99 (0.93–1.05) | 0.747 | 1.01 (0.95–1.08) | 0.644 |
| **Geographic Region** |  |  |  |  |
| Northeast (Ref) | 1.00 |  | 1.00 |  |
| Midwest | 0.98 (0.71–1.34) | 0.880 | 0.71 (0.52–0.98) | 0.037 |
| South | 0.92 (0.75–1.13) | 0.437 | 0.99 (0.82–1.21) | 0.958 |
| West | 0.81 (0.66–0.99) | 0.036 | 0.80 (0.64–0.98) | 0.033 |
| **Hospital Bed Size** |  |  |  |  |
| Large ≥400 (Ref) | 1.00 |  | 1.00 |  |
| Medium (200-399) | 0.83 (0.63–1.10) | 0.196 | 1.00 (0.78–1.27) | 0.981 |
|  | OR (95% CI) | P value | aOR (95% CI) | P value |
| Small (<200) | 0.55 (0.41–0.74) | <0.001 | 0.73 (0.57–0.94) | 0.015 |
| **Teaching Hospital** |  |  |  |  |
| Non-Teaching (Ref) | 1.00 |  | 1.00 |  |
| Teaching | 1.19 (0.99–1.44) | 0.069 | 1.05 (0.89–1.24) | 0.566 |
| **NCI Designation** |  |  |  |  |
| Non-NCI (Ref) | 1.00 |  | 1.00 |  |
| Clinical Cancer Center | 2.40 (1.43–4.04) | <0.001 | 1.95 (1.09–3.50) | 0.025 |
| Comprehensive Cancer Center | 2.23 (1.57–3.16) | <0.001 | 1.88 (1.28–2.76) | 0.001 |
| **CoC Accreditation** |  |  |  |  |
| Non-CoC (Ref) | 1.00 |  | 1.00 |  |
| CoC-Accredited | 1.31 (1.11–1.55) | 0.001 | 1.04 (0.88–1.24) | 0.646 |

Multivariable model adjusted for all variables shown, with missing indicators for Gleason score, PSA, stage, SES quintile, teaching status, and NCI/CoC designation (not shown). GEE models use an exchangeable working correlation and empirical standard errors to account for hospital-level clustering. aOR, adjusted odds ratio; CI, confidence interval; CoC, Commission on Cancer; GEE, generalized estimating equations; NCI, National Cancer Institute; NH, Non-Hispanic; OR, odds ratio; PSA, prostate-specific antigen; Ref, reference; SES, socioeconomic status.

A higher comorbidity burden was associated with lower odds of ODD. Patients with CCI 1 (aOR 0.86, 95% CI 0.82–0.90) and CCI 2+ (aOR 0.64, 95% CI 0.60–0.69) had lower odds of ODD compared with CCI 0 patients. Patients with Gleason 8–10 scores had lower odds of ODD than those with a Gleason score ≤6 (aOR 0.91, 95% CI 0.84–0.99), while Gleason 7 showed no significant difference (aOR 0.99, 95% CI 0.93–1.06). Compared with those with PSA ≤10 ng/mL, those with PSA 10–20 ng/mL had lower odds of ODD (aOR 0.93, 95% CI 0.86–0.99), and those with PSA >20 ng/mL had lower odds still (aOR 0.80, 95% CI 0.70–0.91).

There was evidence of a socioeconomic gradient in ODD. Patients in the lowest compared with the highest Yost quintile had 14% lower odds of ODD (aOR 0.86, 95% CI 0.79–0.94).

Unmarried patients had 21% lower odds of ODD than married patients (aOR 0.79, 95% CI 0.73– 0.85). Patients treated at small hospitals (<200 beds) compared with large hospitals (≥400 beds) had 27% lower odds of ODD (aOR 0.73, 95% CI 0.57–0.94).

### Geographic and temporal variation in one-day discharge

Regional ODD rates showed modest variation: Northeast, 56.7%; South, 50.8%; West, 51.3%; and Midwest, 49.1% (Table 1). After adjusting for patient demographics, comorbidities, cancer characteristics, hospital resources, and temporal trends, geographic differences persisted in the multivariable GEE analysis (Fig 2). Patients from the Midwest (aOR 0.71, 95% CI 0.52–0.98) and the West (aOR 0.80, 95% CI 0.64–0.98) had significantly lower odds of ODD compared with those in the Northeast, while the South showed no significant difference (aOR 0.99, 95% CI 0.82–1.21).

ODD increased over the study period, consistent with higher nationwide adoption of perioperative care innovations and increased utilization of robotic surgery relative to open surgery (Fig 3). Patients treated during the late period (2014–2019) had 31% higher odds of ODD than those treated in the early period (2008–2013) (aOR 1.31, 95% CI 1.20–1.43). The overall annual ODD rate increased from 38.4% in 2008 to 59.0% in 2017. In a sensitivity analysis excluding variables with high missingness (Gleason score, PSA, and cancer stage), regional odds ratios were attenuated toward the null (Midwest aOR 0.81, 95% CI 0.59–1.11; South aOR 1.01, 95% CI 0.83–1.23; West aOR 0.89, 95% CI 0.73–1.09), with the Midwest and West no longer reaching statistical significance, suggesting that missingness in clinical variables may contribute some residual confounding to regional estimates (S1 Fig).

**Fig 3.**
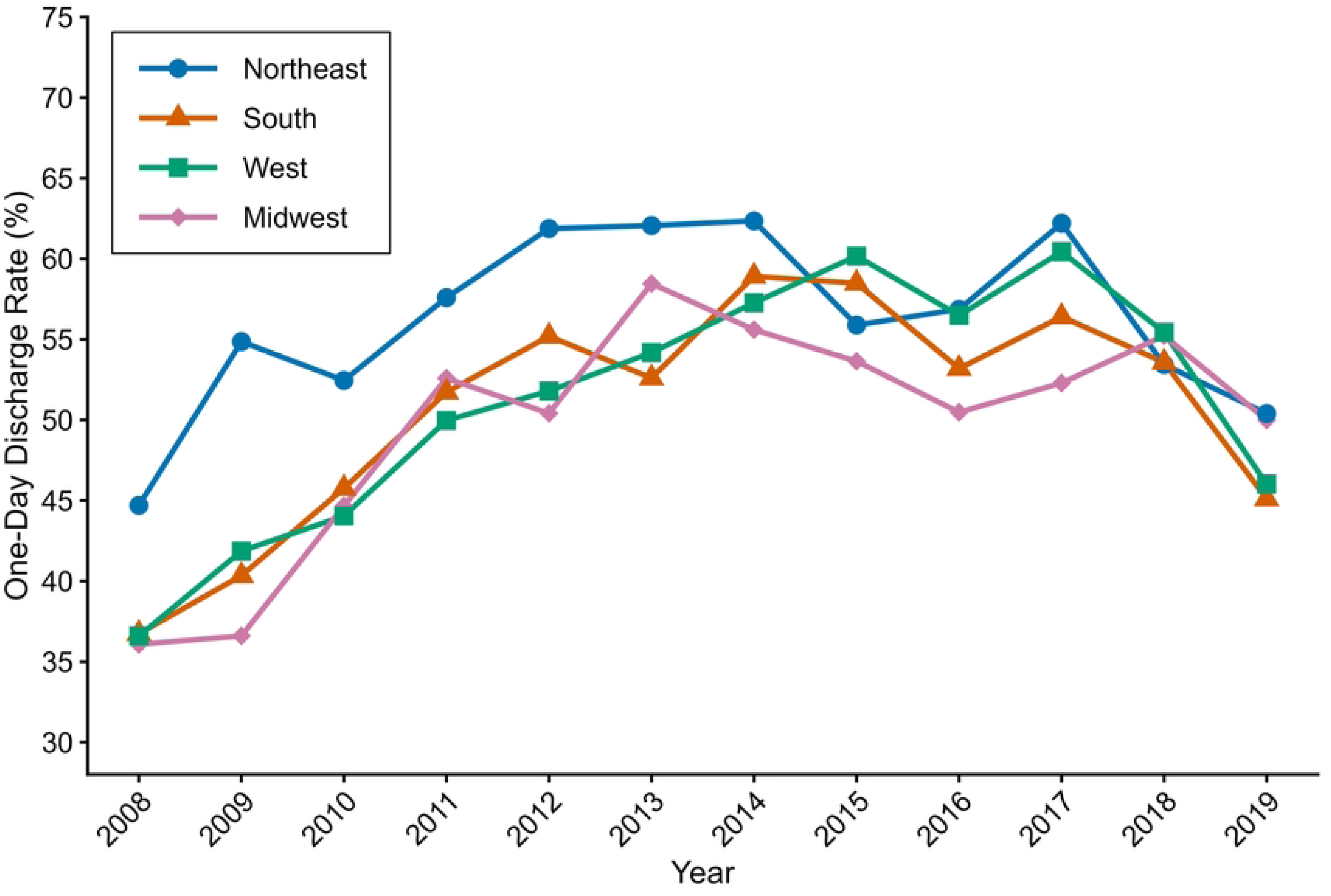
Annual one-day discharge rates by geographic region, 2008–2019. One-day discharge rates are plotted annually for each SEER census region. SEER, Surveillance, Epidemiology, and End Results.

## Discussion

This population-based analysis demonstrates that ODD following radical prostatectomy has increased substantially over the past decade, with more than half of patients now discharged on postoperative day 1. The robotic surgical approach was the strongest predictor of ODD, followed by treatment at NCI-designated cancer centers and the later treatment era. Geographic variation was observed but attenuated in sensitivity analyses, suggesting that regional differences reflect institutional resources rather than independent practice patterns. Persistent racial and socioeconomic gradients in ODD remained after adjustment for clinical- and hospital-level factors.

During our study period, robotic platforms were adopted broadly across the United States despite limited comparative effectiveness data [2,12,13]. Geographic variation initially appeared significant, with the Midwest and West demonstrating lower adjusted odds than the Northeast. However, these associations were attenuated in sensitivity analyses excluding Gleason score, PSA, and cancer stage, suggesting that these clinical variables may influence regional estimates. Given the high rates of missingness in these variables, possible residual confounding due to missing data should be acknowledged, although the overall conclusions from the main model remained unchanged (S1 Fig).

Smaller hospital size was associated with lower odds of ODD, while NCI-designated cancer centers demonstrated higher discharge rates, likely reflecting specialized expertise, enhanced care coordination, and institutional commitment to perioperative innovation [12]. These findings highlight opportunities for quality improvement at lower-volume centers rather than reflecting inherent quality deficiencies.

Previous studies have shown that Black men face disproportionate prostate cancer incidence and mortality and that standardizing access in equal-access systems can reduce racial differences in surgical outcomes [19–24]. In our study, racial and socioeconomic differences in ODD persisted after adjustment for clinical, demographic, and hospital-level factors. While these patterns may reflect barriers to perioperative care, they may also reflect appropriate clinical decision making. For example, patients without reliable support at home may benefit from a longer monitored recovery stay. Future studies should explore whether ODD is equally safe and beneficial across all patient populations before longer hospital stays are characterized as a negative outcome. This study has several limitations that merit consideration. Administrative data sources lack granular details of surgical complexity and postoperative complications that may influence discharge timing. The inability to distinguish specific surgical approaches beyond robotic versus non-robotic in MEDPAR data limits technique-specific analysis. These results may not be generalizable to younger or commercially insured populations. Notably, 50.5% of Gleason scores and 45.9% of PSA values were missing, which may limit the precision of estimates for these variables; however, missing indicators were included in the models to retain the full analytic sample. Sensitivity analyses excluding Gleason score, PSA, and cancer stage showed that regional estimates were attenuated, suggesting possible residual confounding from data missingness, although the direction and magnitude of the primary findings were largely preserved. We lacked information about specific clinical practice changes that may have driven the observed increases in ODD [14].

## Conclusions

This population-based study demonstrated that ODD after radical prostatectomy has become the predominant practice pattern among Medicare beneficiaries, driven primarily by temporal adoption trends and robotic surgery rather than geographic factors. While racial and socioeconomic differences in discharge patterns persisted, future work is needed to determine whether early discharge is universally beneficial or whether some patients may be better served by a longer monitored recovery.

## Data Availability

The data underlying this study are third-party data from the SEER-Medicare linked database, obtained from the U.S. National Cancer Institute (NCI) under a data use agreement. The authors are not permitted to share these data publicly because they contain protected health information governed by NCI and the Centers for Medicare & Medicaid Services. Other researchers may request access to identical SEER-Medicare data through the NCI Healthcare Delivery Research Program (https://healthcaredelivery.cancer.gov/seermedicare/obtain/), subject to approval of a data request and execution of a data use agreement. The authors did not have any special access privileges that others would not have. SAS code used for cohort construction and analysis is available from the corresponding author on request.

## Acknowledgments

The collection of cancer incidence data used in this study was supported by the California Department of Public Health pursuant to California Health and Safety Code Section 103885; Centers for Disease Control and Prevention’s (CDC) National Program of Cancer Registries, under cooperative agreement 1NU58DP007156; the National Cancer Institute’s Surveillance, Epidemiology and End Results Program under contract HHSN261201800032I awarded to the University of California, San Francisco, contract HHSN261201800015I awarded to the University of Southern California, and contract HHSN261201800009I awarded to the Public Health Institute. The ideas and opinions expressed herein are those of the author(s) and do not necessarily reflect the opinions of the State of California, Department of Public Health, the National Cancer Institute, and the Centers for Disease Control and Prevention or their Contractors and Subcontractors. This study used the linked SEER-Medicare database. The interpretation and reporting of these data are the sole responsibility of the authors. The authors acknowledge the efforts of the National Cancer Institute; Information Management Services (IMS), Inc.; and the Surveillance, Epidemiology, and End Results (SEER) Program tumor registries in the creation of the SEER-Medicare database.

## Supporting information

**S1 Checklist. STROBE checklist for cohort studies.**

**S1 Table. Procedure codes used for cohort identification.** Radical prostatectomy and robotic-assisted surgery codes used to identify the study cohort from MEDPAR inpatient claims (SEER-Medicare 2008–2019).

**S2 Table. ICD diagnosis codes used for the Charlson Comorbidity Index.** Comorbidities were identified using the NCI Comorbidity Macro applied to Medicare claims in the 12 months prior to the month of radical prostatectomy admission.

**S1 Fig. Sensitivity analysis excluding Gleason score, PSA, and cancer stage.** Forest plot of adjusted odds ratios and 95% confidence intervals for one-day discharge from the multivariable GEE model re-estimated without Gleason score, PSA, and cancer stage. GEE, generalized estimating equations; PSA, prostate-specific antigen.

